# KNOWLEDGE AND RISK PERCEPTION OF NIGERIANS TOWARDS THE CORONAVIRUS DISEASE (COVID-19)

**DOI:** 10.1101/2021.07.30.21261351

**Authors:** Bolaji Felicia Udomah, Uriel Oludare Ashaolu, Charles Oluwatemitope Olomofe, Olufunke Folasade Dada, Victor Kehinde Soyemi, Yetunde Bolatito Aremu-Kasumu, Chikezie John Ochieze, Ayodele Olusola Adeyemi, Adeyinka Olabisi Owolabi, Martin Chukwudum Igbokwe, Emmanuel Eziashi Ajumuka, Kehinde Williams Ologunde, Gbenga Omotade Popoola, Olumuyiwa Elijah Ariyo, Olaniyi Bamidele Fayemi

**Author notes:** Corresponding author:Charles Oluwatemitope Olomofe.

## Abstract

**Background:** The Coronavirus Disease 2019 (COVID-19) is far from over, although appreciable progress has been made to limit the devastating effects of the pandemic across the globe. Adequate knowledge and risk perception is a critical assessment that is required to ensure proper preventive measures. This study assessed these among Nigerians.

**Methods:** The study was a cross-sectional assessment of 776 consenting Nigerian adults that were distributed across the 6 geo-political zones and the Federal Capital Territory. Online pre-tested, semi-structured questionnaire were used to obtain the socio-demographic data and assessed the knowledge and risk perception of the participants to COVID-19. The knowledge of COVID-19 was assessed based on the number of accurate responses given in comparison to average scores. Chi-square analysis was computed to analysis the association between socio-demographic characteristics and knowledge of COVID-19 and risk perception. Data analysis was done using SPSS version 21, the level of significance was set at value p<0.05 at 95% confidence interval.

**Results:** Majority of the participants were male 451 (58.1%), there was a good knowledge of COVID-19 among 90.3% of respondents with 57% having positive risk perception. There was a statistically significant relationship between good knowledge and positive risk perception of COVID-19 (p < 0.001). Annual income (p =0.012) and the perception that “vaccines are good” significantly predict positive risk perception of COVID-19 among the respondents.

**Conclusion:** A good knowledge of COVID-19 and vaccination against the virus were the two most important factors that determined risk perception among the population. This may be because of the widespread advocacy, and it portends a good omen at combating COVID-19 menace.

## BACKGROUND

The novel coronavirus (Severe Acute Respiratory Syndrome Coronavirus 2, SARS-CoV-2) was first reported in Wuhan city in China, and as of December 2020, 191 countries of the world have been affected by it.^1^ As of 16th January 2021, there were over 90 million confirmed cases worldwide with more than 2 million mortality while Nigeria had corresponding figures of 105, 478, and 1,405 respectively.^2,3^ Coronaviruses are a large family of viruses that are known to cause illnesses ranging from the common cold to more severe diseases such as Middle East Respiratory Syndrome (MERS) and Severe Acute Respiratory Syndrome (SARS-CoV2).^4^ The clinical symptoms of Coronavirus Disease 2019 (COVID-19) include fever (which is the most common), cough, fatigue, malaise, and shortness of breath,^5^ and these symptoms are more severe amongst the elderly and those with underlying chronic medical conditions such as diabetes, chronic heart, and chronic lung diseases.^5,6^

Many countries, including Nigeria experienced a second wave of the viral infection. The total number of COVID-19 infected persons is almost reaching 100,000.^7^ This becomes complicated because of new variants of the virus has also been discovered in Nigeria, which was first isolated in the United Kingdom (UK), said to be about 56% more transmissible, with a large number of mutation and virulence.^8^ Since the beginning of December there has been an astronomical increase in the number of new cases and deaths recorded on daily basis.^9^ Experts have linked this with non-adherence to the infection control recommendations.^10^ These recommendations are focused on disease prevention and control measures to minimise the spread as well as the burden on the healthcare system.^11^ These measures include frequent hand hygiene using soap and water or an alcohol-based sanitizer, keeping a physical distance of about 6 feet from others, quarantine for those who are exposed, use of face masks, and avoiding the touch of the face with unwashed hands.^12^ However, many Nigerians still do not adhere to these preventive measures. Good knowledge, and risk perception which refers to the way the diseases is understood/interpreted,^13^ is important in pre-empting the way a person would react to an occurrence of the infection in their own lives as well as the lives of others. An understanding of the perception of people regarding COVID-19 is crucial because perception can act as a trigger for precautionary action, engagement in preventive health behaviour and reporting of suspected cases of COVID-19.^14^ Knowledge and perception of risk is influenced by both individual and societal factors which are based on experiences, beliefs, attitudes, social, cultural and institutional processes.^13^ Risk perception is also largely mediated by the information and knowledge that an individual has. Bhagavathula et al,^15^ in their study assessed the awareness of COVID-19 among health care workers reported that knowledge of transmission (60%) and disease onset (64%) was poor among the health care workers.

The Nigerian government, non-governmental organizations (NGOs), and civil societies are using various avenues to bridge the knowledge gap by educating and enlightening the populace on COVID-19 and how to prevent its spread. One of such measures is the collaboration by the Nigerian Red Cross Society with tricycle riders in Maiduguri where they are providing stickers for them to use on their tricycles across the city.^16^ Also, several media strategies such as campaigns on social media platforms, press releases, radio jingles and drama skits in local dialects are being used.^17^ A survey conducted among Egyptians shows many had good knowledge of the disease but are not willing to practice the preventive measures.^18^ Anecdotal evidence suggests poor knowledge and poor perception of risk of the disease amongst the Nigerian population could be responsible for poor adherence to these preventive measures.^19^ Yet there is dearth of studies evaluating the knowledge and perception of the people about the on-going COVID 19 pandemic among Nigerians. The elites in governance, policymakers and opinion leaders have been reported to have questioned the need for social distancing, while others have cited political, economic, and financial gains as the drive(s) for the fight against the infection.^20^ Similarly, a cross-section of people on the streets also gives overwhelming evidence that Nigerians are largely ignoring COVID-19 safety protocols.^21^ Therefore, there is a need to assess the level of knowledge of Nigerians about COVID-19 and their perceived risk of contracting the disease. The outcome of this research may reveal if their knowledge and risk perception of contracting COVID-19 is adequate or not and inform necessary action to improve the knowledge and risk perception targeted at proper adherence to preventive measures.

## METHODS

This was a cross sectional study carried out in Nigeria, the most populated country in Africa with an estimated population of about 200 million and a total land area of 910,770 Km2 (351,650 sq. miles).^22^ Nigeria has six geopolitical zones (Southwest, Southeast, Southsouth, Northwest, Northeast, Northcentral) with 36 states. Ethical approval was gotten from the health research ethics committee of the Federal Medical Centre Gusau, Zamfara State.

This was an on-line study, conducted using a pre-tested, semi-structured questionnaire and included Nigerian adults above the age of 18 years who consented to participate in the study. The minimum sample size was determined to be 409 at confidence level of 95% and based on proportion of people with good knowledge of 39% in a previous study^23^ and a 5% margin of error.

The questionnaire was adapted from several published literature^23–25^ and covered three sections A-C that is, Socio-demographic characteristics of respondents, respondent’s knowledge of COVID 19, respondents risk perception of COVID 19. Pre-test of the questionnaire was done on 10% of the respondents each at six different states from the six geopolitical zones and were not included in the study. The pretested questionnaires with participants’ information sheet were circulated extensively online.

Data was analyzed using SPSS version 21. The knowledge of COVID 19 among respondents was scored based on the number of accurate responses given. The number of correct responses were compared with the average score. Participants whose score equalled or was above the average were categorized as having good knowledge while those who scored below the average were categorized as having poor knowledge. Chi square analysis was computed to test for association between sociodemographic characteristics and knowledge of COVID 19 among respondents. The level of significance was predetermined at a p-value of less than 0.05 at 95% confidence level.

## RESULTS

A total of 776 participants completed the survey. Most were within the ages of 36-45 years (43.9%), with 58.1% males and 40.9% females. The majority (53.2%) had tertiary education with 7.7% of the respondents being artisans, 8.6% being teachers and 25% being health care workers. Most respondents (26.7%) preferred not to say their annual income with 17.5% earning less than 500,000 Naira /annum. Zonal representation of respondents (state of origin and place of residence) revealed that most of the respondents were from the southwest zone of the country (47.4% and 46% respectively). Most respondents’ households were made up of 1-4 persons (48.3%) while 5.7% of the respondents had more than 8 persons per household (**Table 1**).

**Table 1:** Socio-demographic characteristics of Respondents.

| **Variables** | **Frequency (*n* = 776)** | **Percent (%)** |
| --- | --- | --- |
| **Age group (years)** |  |  |
| *18 – 25* | 93 | 12.0 |
| *26 – 35* | 241 | 31.1 |
| *36 – 45* | 341 | 43.9 |
| *46 – 55* | 73 | 9.4 |
| *> 55* | 28 | 3.6 |
| **Gender** |  |  |
| *Male* | 451 | 58.1 |
| *Female* | 317 | 40.9 |
| *Prefer not to say* | 8 | 1.0 |
| **Marital Status** |  |  |
| *Single* | 214 | 27.6 |
| *Married* | 489 | 63.0 |
| *Separated/ Divorced* | 15 | 1.9 |
| *Widowed/ Widower* | 7 | 0.9 |
| *In a relationship* | 45 | 5.8 |
| *Undisclosed* | 6 | 0.8 |
| **Religion** |  |  |
| *None* | 9 | 1.2 |
| *Christianity* | 616 | 79.4 |
| *Islam* | 147 | 18.9 |
| *Others* | 4 | 0.5 |
| **Level of Education** |  |  |
| *Secondary school or less* | 36 | 4.6 |
| *Tertiary* | 413 | 53.2 |
| *Postgraduate* | 327 | 42.1 |
| **Occupation** |  |  |
| *Unemployed* | 60 | 7.7 |
| *Trader* | 22 | 2.8 |
| *Teacher* | 67 | 8.6 |
| *Artisan* | 17 | 2.2 |
| *Healthcare worker* | 194 | 25.0 |
| *Information technology* | 43 | 5.5 |
| *Financial services* | 45 | 5.8 |
| *Security agent* | 12 | 1.5 |
| *Agricultural services* | 19 | 2.4 |
| *Others* | 297 | 38.3 |
| **Estimated Income of Caregiver**<br>**Per Annum (Naira)** |  |  |
| *< 500,000* | 136 | 17.5 |
| *500,000 -1 million* | 120 | 15.5 |
| *1 – 2 million* | 117 | 15.1 |
| *2 – 3 million* | 54 | 7.0 |
| *3 – 4 million* | 37 | 4.8 |
| *> 4 million* | 105 | 13.5 |
| *I prefer not to say* | 207 | 26.7 |
| **State Origin (Zone)** |  |  |
| *North-Central* | 90 | 11.6 |
| *North-East* | 46 | 5.9 |
| *North-West* | 49 | 6.3 |
| *South-East* | 119 | 15.3 |
| *South-South* | 94 | 12.1 |
| *South-West* | 368 | 47.4 |
| *Prefer not to say* | 10 | 1.3 |
| **Place of residence (Zone)** |  |  |
| *North-Central* | 174 | 22.4 |
| *North-East* | 24 | 3.1 |
| *North-West* | 103 | 13.3 |
| *South-East* | 32 | 4.1 |
| *South-South* | 40 | 5.2 |
| *South-West* | 357 | 46.0 |
| *Prefer not to say* | 46 | 5.9 |
| **No person in household** |  |  |
| 1 – 4 | 375 | 48.3 |
| 5 – 8 | 357 | 46.0 |
| > 8 | 44 | 5.7 |

Overall, majority (90.3%) of the respondents had a good knowledge of COVID 19 while the remaining 9.7% had poor knowledge (**Figure 1)**. Most of the respondents (71.8%) reported multiple sources of awareness of COVID 19 including the mass media, immediate community, and hospital-related sources. About a quarter of the respondents reported mass media as their only source of awareness (**Table 2)**.

**Fig 1:**
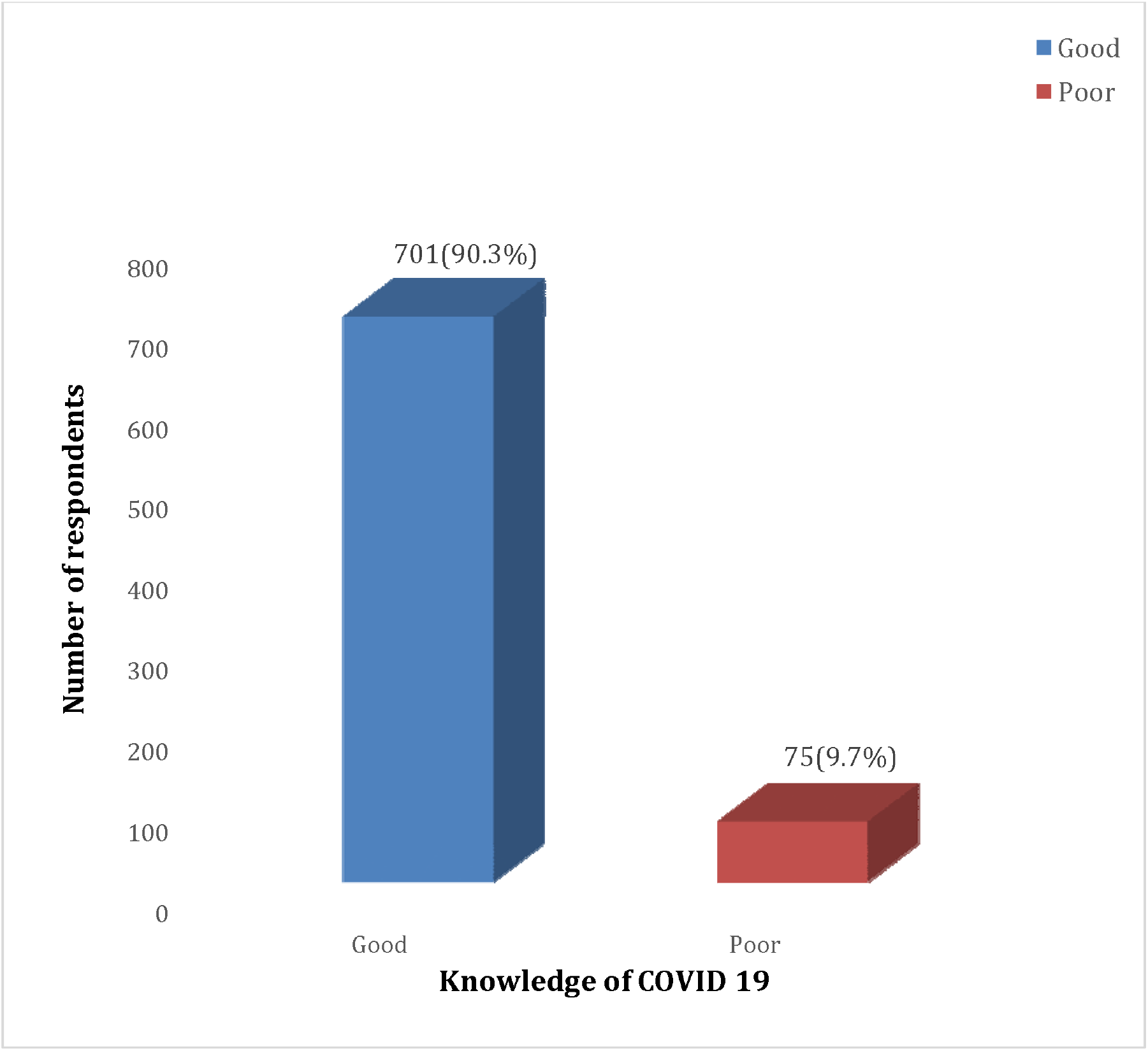
Overall knowledge of COVID-19 among respondents.

**Table 2:** Respondents’ source of awareness of COVID 19.

| Source of awareness of COVID-19 | Frequency (*n* = 776) | Percent (%) |
| --- | --- | --- |
| *Radio/television/newspaper/social media* | 198 | 25.5 |
| *Family/friends/neighbors* | 5 | 0.6 |
| *Healthcare workers/hospital/pharmacy* | 12 | 1.5 |
| *All the above* | 557 | 71.8 |
| *Others (Mosque, Church etc.)* | 4 | 0.5 |

Univariate distribution of the risk perception of respondents revealed that almost all the respondents (98%) agreed that COVID 19 is a global disease and can affect both adults and children. In addition, most of them (94%) agreed that as individuals they were doing a lot to protect their family, and majority also agreed that people can die or recover from the disease. Surprisingly, many of the respondents were indifferent towards government and health system response in tackling the disease (30.4% and 31.7% respectively) and majority were also either neutral or disagreed that COVID 19 is a threat to humanity. More than half of the respondents disagreed that the disease affects the elderly with about 30% indifferent about going to the hospital when ill **(Table 3)**.

**Table 3:** Risk Perception on COVID 19 among Respondents.

| Risk Perception | SA<br>n (%) | A<br>n (%) | N<br>n (%) | DA<br>n (%) | SD<br>n (%) | DK<br>n (%) |
| --- | --- | --- | --- | --- | --- | --- |
| *COVID-19 is a global disease* | 711(91.6) | 50(6.4) | 7(0.9) | 2(0.3) | 2(0.3) | 4(0.5) |
| *The disease can affect both adults and children* | 662(85.3) | 98(12.6) | 7(0.9) | 1(0.1) | 1(0.1) | 7(0.9) |
| *I am doing enough to protect myself and family* | 480(61.9) | 250(32.2) | 31(4.0) | 5(0.6) | 1(0.1) | 9(1.2) |
| *Government is doing enough to protect myself and my family* | 89(11.5) | 201(25.9) | 236(30.4) | 144(18.6) | 90(11.6) | 16(2.1) |
| *Our healthcare system can handle the disease* | 71(9.1) | 150(19.3) | 246(31.7) | 186(24.0) | 103(13.3) | 20(2.6) |
| *The disease is not a threat to human race* | 60(7.7) | 150(19.3) | 182(23.5) | 186(24.0) | 164(21.1) | 34(4.4) |
| *People can die from the disease* | 624(80.4) | 112(14.4) | 16(2.1) | 6(0.8) | 6(0.8) | 12(1.5) |
| *People can recover after having the disease* | 544(70.1) | 194(25.0) | 23(3.0) | 2(0.3) | 4(0.5) | 9(1.2) |
| *It is only old people that die from the disease* | 22(2.8) | 93(12.0) | 182(23.5) | 251(32.3) | 174(22.4) | 54(7.0) |
| *Even if am ill now I would not go to the hospital because of the risk of contracting the COVID infection* | 98(12.6) | 181(23.3) | 230(29.6) | 145(18.7) | 90(11.6) | 32(4.1) |

**A = Agree, SA = Strongly Agree, N = Neutral, D = Disagree, SD = Strongly Disagree DK = Don't Know**

Overall, 57% of the respondents had a positive risk perception of COVID-19 while 43% had a negative risk perception **(Figure 2)**. Sociodemographic variables such as age, marital status and occupation of the respondents were found to significantly influence the knowledge of the respondents on COVID 19 **(Table 4a and 4b)**.

**Fig 2:**
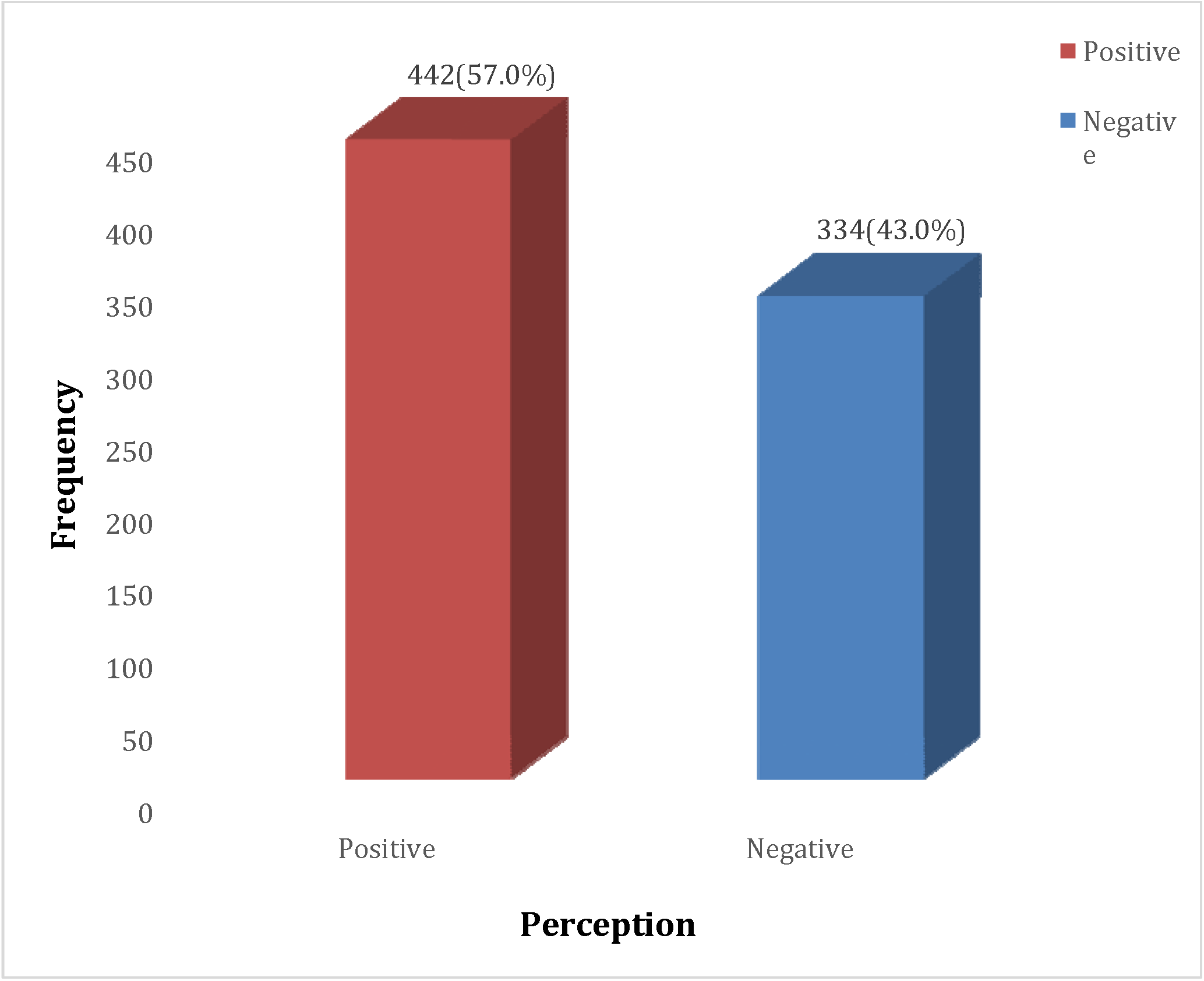
Overall risk perception of COVID-19 among respondents.

**Table 4a:** Association between Sociodemographic Characteristics and Knowledge of COVID 19.

| Socio-Demographic Characteristics | Knowledge | | | $\chi^2$ | *p*-value |
| --- | --- | --- | --- | --- | --- |
|  | Good<br>n (%) | Poor<br>n (%) | Total<br>N (%) |  |  |
| **Age (years)** |  |  |  |  |  |
| 18 – 25 | 87 (93.5) | 6 (6.5) | 93 | 12.674 | **0.013\*** |
| 26 – 35 | 219 (90.9) | 22 (9.1) | 242 |  |  |
| 36 – 45 | 309 (90.6) | 32 (9.4) | 341 |  |  |
| 46 – 55 | 66 (90.4) | 7 (9.6) | 73 |  |  |
| > 55 | 20 (71.4) | 8 (28.6) | 28 |  |  |
| **Gender** |  |  |  |  |  |
| Male | 411 (91.1) | 40 (8.9) | 451 | 0.514 | 0.473 |
| Female | 284 (89.6) | 33 (10.4) | 317 |  |  |
| **Level education** |  |  |  |  |  |
| Secondary school or less | 32 (88.9) | 4 (11.1) | 36 | 2.640 | 0.267 |
| Tertiary | 367 (88.9) | 46 (11.1) | 413 |  |  |
| Postgraduate | 302 (92.4) | 25 (7.6) | 327 |  |  |
| **Marital status** |  |  |  |  |  |
| Single | 189 (88.3) | 25 (11.7) | 214 | 11.241<sup>F</sup> | **0.017\*** |
| Married | 446 (91.2) | 43 (8.8) | 489 |  |  |
| Separated/ Divorced | 13 (86.7) | 2 (13.3) | 15 |  |  |
| Widowed/ Widower | 4 (57.1) | 3 (42.9) | 7 |  |  |
| In a relationship | 44 (97.8) | 1 (2.2) | 45 |  |  |
| **Number person in household** |  |  |  |  |  |
| 1 – 4 | 335 (89.3) | 40 (10.7) | 375 | 1.227 | 0.541 |
| 5 – 8 | 327 (91.6) | 30 (8.4) | 357 |  |  |
| > 8 | 39 (88.6) | 5 (11.4) | 44 |  |  |
| **State origin zone** |  |  |  |  |  |
| North-Central | 78 (86.7) | 12 (13.3) | 90 | 10.440 | 0.064 |
| North-East | 40 (87.0) | 6 (13.0) | 46 |  |  |
| North-West | 43 (87.8) | 6 (12.2) | 49 |  |  |
| South-East | 108 (90.8) | 11 (9.2) | 119 |  |  |
| South-South | 80 (85.1) | 14 (14.9) | 94 |  |  |
| South-West | 345 (93.8) | 23 (6.3) | 368 |  |  |
| **Place of residence** |  |  |  |  |  |
| North-Central | 151 (86.8) | 23 (13.2) | 174 | 10.015<sup>F</sup> | 0.061 |
| North-East | 21 (87.5) | 3 (12.5) | 24 |  |  |
| North-West | 95 (92.2) | 8 (7.8) | 103 |  |  |
| South-East | 30 (93.8) | 2 (6.3) | 32 |  |  |
| South-South | 33 (82.5) | 7 (17.5) | 40 |  |  |
| South-West | 333 (93.3) | 24 (6.7) | 357 |  |  |
| **Religion** |  |  |  |  |  |
| None | 8 (88.9) | 1 (11.1) | 9 | 1.777 | 0.606 |
| Christianity | 560 (90.9) | 56 (9.1) | 616 |  |  |
| Islam | 129 (87.8) | 18 (12.2) | 147 |  |  |
| Others | 4 (100.0) | 0 (0.0) | 4 |  |  |

**Table 4b:**
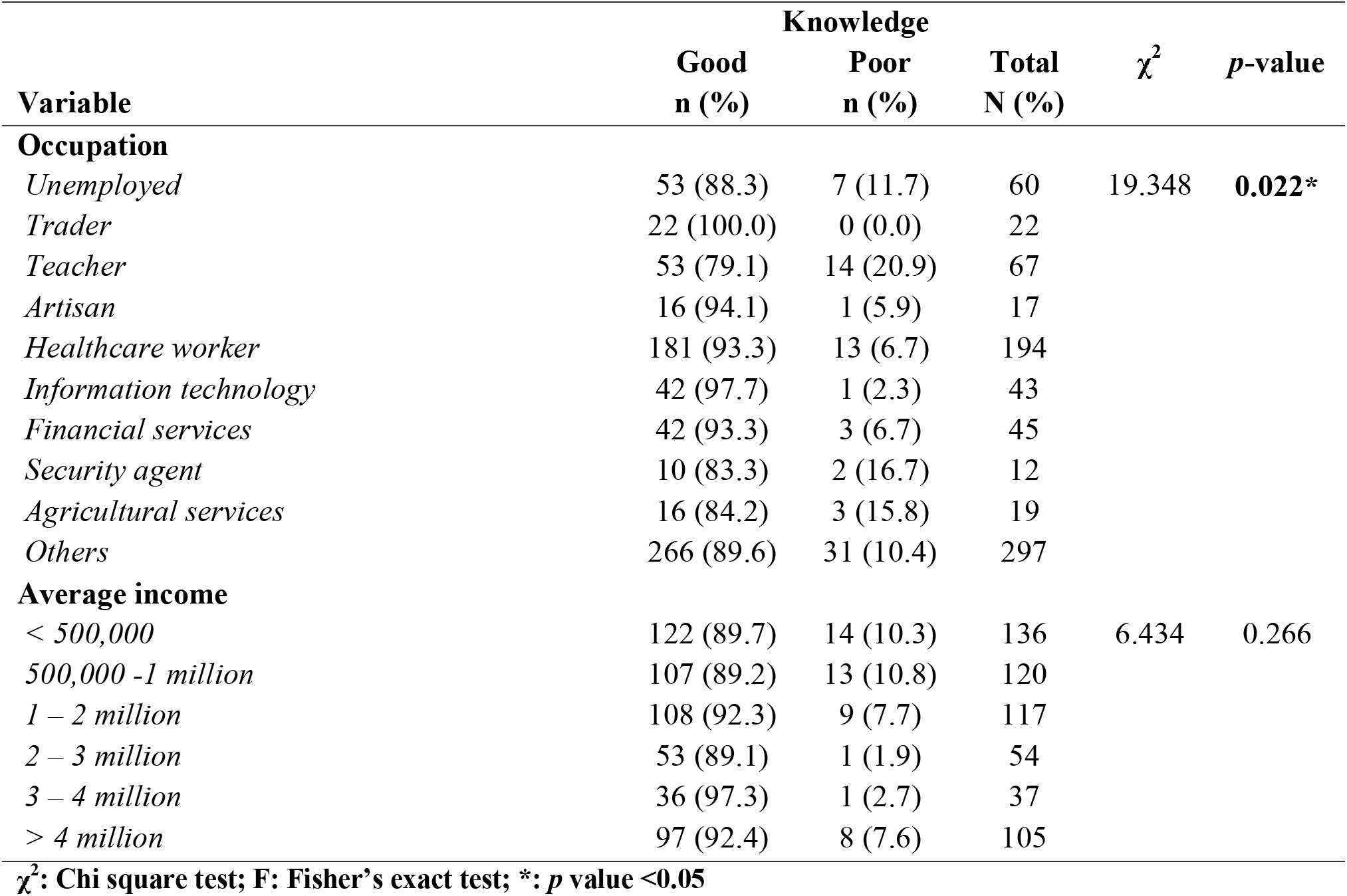
Association between Sociodemographic Characteristics and Knowledge of COVID 19.

Four hundred and eighteen (59.6%) of the seven hundred and one respondents with good knowledge about COVID 19 also had positive perception of COVID 19 risk. Contrarily, only twenty-four (32%) of seventy five subjects with poor COVID 19 knowledge had positive risk perception. Thus, we found a statistically significant relationship between good knowledge and positive risk perception of COVID 19 (p < 0.001). **(Table 5)**

**Table 5:** Association between knowledge of COVID-19 and risk perception of COVID-19.

| Variable | Risk Perception of COVID 19 | | | $\chi^2$ | p-value |
| --- | --- | --- | --- | --- | --- |
|  | Positive<br>n (%) | Negative<br>n (%) | Total<br>N (%) |  |  |
| Knowledge of COVID-19 |  |  |  |  |  |
| Good | 418 (59.6) | 283 (40.4) | 701 | 21.096 | <0.001* |
| Poor | 24 (32.0) | 51 (68.0) | 75 |  |  |

$\chi^2$ : Chi Square test; \*: p value <0.05

Significant predictors of positive risk perception include annual income (*p = 0*.*012, OR = 3*.*676, CI 1*.*324 - 10*.*212*), the perception that ‘vaccines are good’ (*p = 0*.*013, OR = 2*.*122, CI 1*.*175 - 3*.*835*) and good knowledge of COVID 19 (*p = 0*.*019, OR = 2*.*578, CI 1*.*167 - 5*.*695*). Age, level of education, marital status, place of readiness, religion, occupation, readiness to receive the COVID 19 vaccine and history of previous vaccinations did not significantly predict COVID 19 risk perception. **(Table 6)**

**TaBLE 6:**
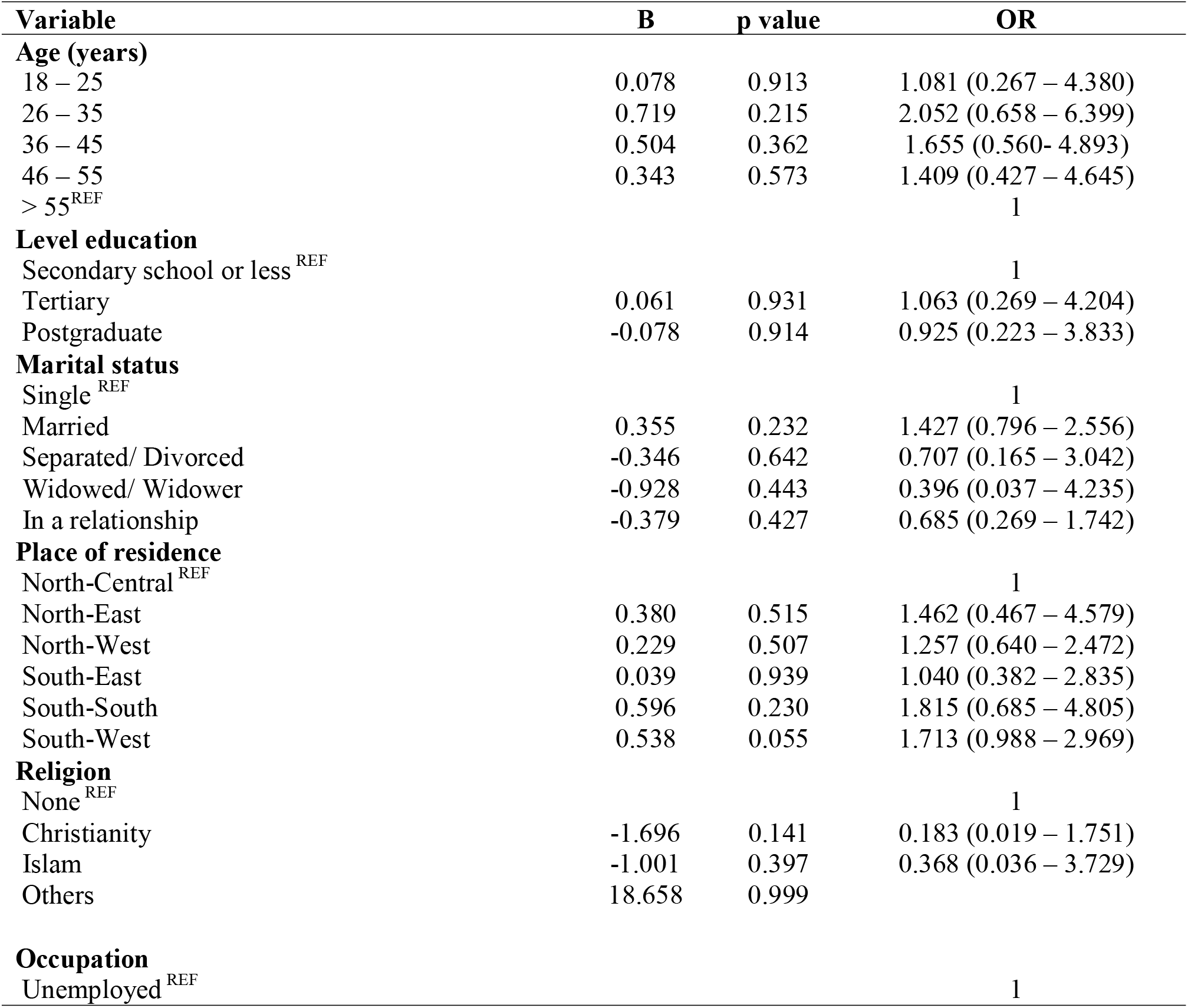
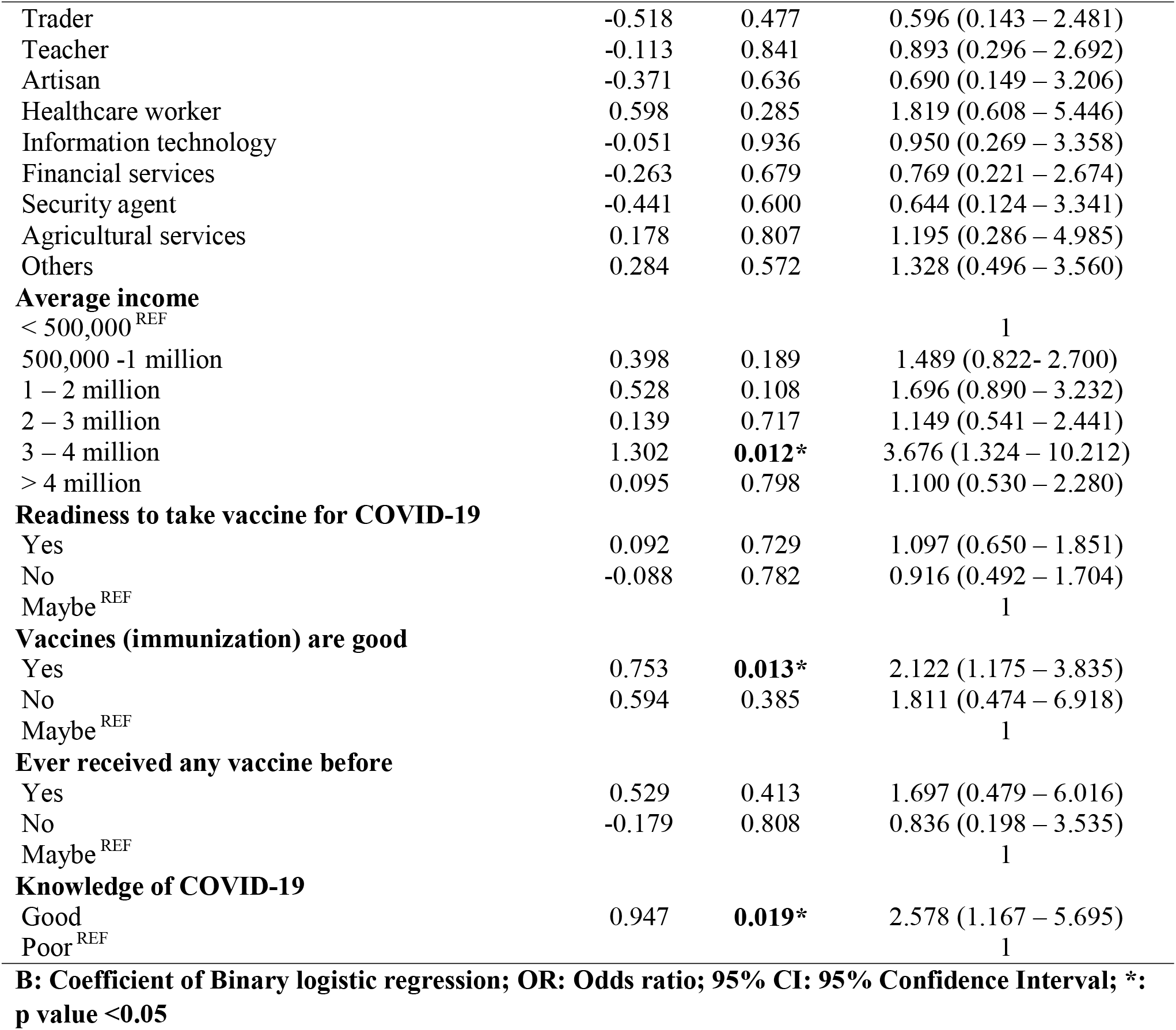
Predictors of positive risk perception of COVID-19.

| Variable | B | p value | OR |
| --- | --- | --- | --- |
| **Age (years)** |  |  |  |
| 18 – 25 | 0.078 | 0.913 | 1.081 (0.267 – 4.380) |
| 26 – 35 | 0.719 | 0.215 | 2.052 (0.658 – 6.399) |
| 36 – 45 | 0.504 | 0.362 | 1.655 (0.560- 4.893) |
| 46 – 55 | 0.343 | 0.573 | 1.409 (0.427 – 4.645) |
| > 55<sup>REF</sup> |  |  | 1 |
| **Level education** |  |  |  |
| Secondary school or less<sup>REF</sup> |  |  | 1 |
| Tertiary | 0.061 | 0.931 | 1.063 (0.269 – 4.204) |
| Postgraduate | -0.078 | 0.914 | 0.925 (0.223 – 3.833) |
| **Marital status** |  |  |  |
| Single<sup>REF</sup> |  |  | 1 |
| Married | 0.355 | 0.232 | 1.427 (0.796 – 2.556) |
| Separated/ Divorced | -0.346 | 0.642 | 0.707 (0.165 – 3.042) |
| Widowed/ Widower | -0.928 | 0.443 | 0.396 (0.037 – 4.235) |
| In a relationship | -0.379 | 0.427 | 0.685 (0.269 – 1.742) |
| **Place of residence** |  |  |  |
| North-Central<sup>REF</sup> |  |  | 1 |
| North-East | 0.380 | 0.515 | 1.462 (0.467 – 4.579) |
| North-West | 0.229 | 0.507 | 1.257 (0.640 – 2.472) |
| South-East | 0.039 | 0.939 | 1.040 (0.382 – 2.835) |
| South-South | 0.596 | 0.230 | 1.815 (0.685 – 4.805) |
| South-West | 0.538 | 0.055 | 1.713 (0.988 – 2.969) |
| **Religion** |  |  |  |
| None<sup>REF</sup> |  |  | 1 |
| Christianity | -1.696 | 0.141 | 0.183 (0.019 – 1.751) |
| Islam | -1.001 | 0.397 | 0.368 (0.036 – 3.729) |
| Others | 18.658 | 0.999 |  |
| **Occupation** |  |  |  |
| Unemployed<sup>REF</sup> |  |  | 1 |
| Trader | -0.518 | 0.477 | 0.596 (0.143 – 2.481) |
| Teacher | -0.113 | 0.841 | 0.893 (0.296 – 2.692) |
| Artisan | -0.371 | 0.636 | 0.690 (0.149 – 3.206) |
| Healthcare worker | 0.598 | 0.285 | 1.819 (0.608 – 5.446) |
| Information technology | -0.051 | 0.936 | 0.950 (0.269 – 3.358) |
| Financial services | -0.263 | 0.679 | 0.769 (0.221 – 2.674) |
| Security agent | -0.441 | 0.600 | 0.644 (0.124 – 3.341) |
| Agricultural services | 0.178 | 0.807 | 1.195 (0.286 – 4.985) |
| Others | 0.284 | 0.572 | 1.328 (0.496 – 3.560) |
| **Average income** |  |  |  |
| < 500,000<sup>REF</sup> |  |  | 1 |
| 500,000 -1 million | 0.398 | 0.189 | 1.489 (0.822- 2.700) |
| 1 – 2 million | 0.528 | 0.108 | 1.696 (0.890 – 3.232) |
| 2 – 3 million | 0.139 | 0.717 | 1.149 (0.541 – 2.441) |
| 3 – 4 million | 1.302 | **0.012\*** | 3.676 (1.324 – 10.212) |
| > 4 million | 0.095 | 0.798 | 1.100 (0.530 – 2.280) |
| **Readiness to take vaccine for COVID-19** |  |  |  |
| Yes | 0.092 | 0.729 | 1.097 (0.650 – 1.851) |
| No | -0.088 | 0.782 | 0.916 (0.492 – 1.704) |
| Maybe<sup>REF</sup> |  |  | 1 |
| **Vaccines (immunization) are good** |  |  |  |
| Yes | 0.753 | **0.013\*** | 2.122 (1.175 – 3.835) |
| No | 0.594 | 0.385 | 1.811 (0.474 – 6.918) |
| Maybe<sup>REF</sup> |  |  | 1 |
| **Ever received any vaccine before** |  |  |  |
| Yes | 0.529 | 0.413 | 1.697 (0.479 – 6.016) |
| No | -0.179 | 0.808 | 0.836 (0.198 – 3.535) |
| Maybe<sup>REF</sup> |  |  | 1 |
| **Knowledge of COVID-19** |  |  |  |
| Good | 0.947 | **0.019\*** | 2.578 (1.167 – 5.695) |
| Poor<sup>REF</sup> |  |  | 1 |

**B: Coefficient of Binary logistic regression; OR: Odds ratio; 95% CI: 95% Confidence Interval; \*: p value <0.05**

Variables that significantly predicted good knowledge of COVID 19 were respondents between the ages of 18 to 25 years and married respondents (*p = 0*.*008, OR = 7*.*474, CI 1*.*697-32*.*920* and *p = 0*.*024, OR=2*.*173, CI 1*.*105-4*.*273* respectively). The occupation of respondents did not significantly predict good knowledge of COVID 19. **(Table 7)**

**Table 7:** Predictors of good knowledge of COVID-19.

| Variable | B | p value | OR | 95% CI |  |
| --- | --- | --- | --- | --- | --- |
|  |  |  |  | Lower | Upper |
| Age (years) |  |  |  |  |  |
| 18 – 25 | 2.011 | 0.008* | 7.474 | 1.697 | 32.920 |
| 26 – 35 | 1.105 | 0.052 | 3.019 | 0.990 | 9.204 |
| 36 – 45 | 0.852 | 0.101 | 2.345 | 0.847 | 6.494 |
| 46 – 55 | 0.819 | 0.193 | 2.267 | 0.661 | 7.781 |
| > 55<sup>REF</sup> |  |  | 1 |  |  |
| Marital status |  |  |  |  |  |
| Single<sup>REF</sup> |  |  | 1 |  |  |
| Married | 0.776 | 0.024* | 2.173 | 1.105 | 4.273 |
| Separated/ Divorced | 0.380 | 0.648 | 1.463 | 0.286 | 7.475 |
| Widowed/ Widower | -0.191 | 0.837 | 0.826 | 0.134 | 5.104 |
| In a relationship | 1.846 | 0.076 | 6.331 | 0.825 | 48.615 |
| Occupation |  |  |  |  |  |
| Unemployed<sup>REF</sup> |  |  | 1 |  |  |
| Trader | 19.727 | 0.998 | 0.000 |  |  |
| Teacher | -0.288 | 0.620 | 0.750 | 0.241 | 2.339 |
| Artisan | 0.759 | 0.507 | 2.135 | 0.227 | 20.062 |
| Healthcare worker | 0.904 | 0.105 | 2.471 | 0.829 | 7.365 |
| Information technology | 2.021 | 0.073 | 7.545 | 0.830 | 68.549 |
| Financial services | 0.998 | 0.198 | 2.713 | 0.594 | 12.401 |
| Security agent | -0.264 | 0.778 | 0.768 | 0.123 | 4.800 |
| Agricultural services | 0.021 | 0.979 | 1.021 | 0.211 | 4.948 |
| Others | 0.491 | 0.339 | 1.633 | 0.598 | 4.463 |

**B:** Coefficient of Binary logistic regression; **OR:** Odds ratio; **95% CI:** 95% Confidence Interval; **REF:** Reference category; **\***: p value <0.05

## DISCUSSION

In this study, most of the respondents were between the age group 36-45 years and majority were males. This is similar to findings obtained in related online studies on COVID 19 in Nigeria.^26, 27^ This is also reflective of the general demographics of Nigeria which is made up of an expansive population pyramid.^28^ The higher male respondent might also be due to disparity in gender accessibility to internet usage in Nigeria which is currently skewed towards male.^29^ This is particularly so, when you consider a similar study done in south west Nigeria using the traditional methods of sample collection, the study revealed a more male dominated respondent than the female, which is in consonance with the findings of this study.^30^

Three socio demographic variables from the study namely, age, marital status, and occupation had statistically significant association with knowledge of COVID 19. This study revealed that health care workers had good knowledge of COVID 19 compared to other occupations sampled. This is similar to another study in Nigeria that showed that health care workers have good knowledge of COVID 19.^31^ Similar study in the northern Nigeria showed that a quarter of respondents got their information about COVID 19 from health care workers which might also inform the good knowledge of health care workers.^32^ This is not surprising as COVID 19 is a health issue and health care workers have been at the forefront of the management of the disease.

The overall knowledge of our respondents on the outbreak of COVID 19 crisis is high at 90.3%, this is probably due to multiple and modern forms of information dissemination available in Nigeria today. Most of our respondents had information from multiple sources. Available evidence showed that internet penetration seem to be high among young Nigerians and this seems to facilitate health information seeking behaviour.^33^ In this study, social media with other electronics media sources were the highest information sources among Nigerians. This was like the findings from other studies done in Nigeria on COVID-19.^34, 35^

The knowledge of the respondent in this study was higher than that of those in the study on COVID 19 crisis in sub-Saharan Africa by Adesegun et al that reported 78%, but comparable to a similar study done on knowledge, attitude, and practice of COVID 19 outbreak, a population-based study in Iran by Erfani et al with 90% demonstrating good knowledge among Iranian population surveyed in the study.^34, 36^

The younger age among the respondents in Nigeria was a predictor of the good knowledge, which was also noted to be strongly associated with their risk perceptions. In other words, young Nigerians have good knowledge about on-going COVID 19 pandemic and are aware of the risk that this poses to the health in general. These findings were similar to the study findings of Shamir et al which showed that information source was significantly determined by socio-demographic characteristics among young Americans and was also associated with both knowledge and beliefs about the pandemic in the United States.^37^

In this study, the risk perceptions associated with COVID 19 diseases was found to be strongly linked to the knowledge and understanding of the disease as a global burden, which could affect both the young and old, as well as being fatal. The pattern seen in this study has further validated reports from other studies on risk perception on COVID 19 which revealed a significant variability in risk perception seen across the globe.^38,39^ Despite the burden COVID 19 poses globally, more than half of the population in this study are indifferent and do not believe that the government is doing enough to protect the general populace from contracting the virus and this is in consonance with a similar study on COVID 19.^40^ This failure of trust in the government’s effort may account for why majority of our respondents don’t rely or patronize government’s health facility as noted in this study, despite government efforts in providing affordable health care to her citizenry. This is also in tandem with a study done in Nigeria which revealed similar findings.^40^

This study also showed that there is a significant association between the amount of knowledge possessed on the disease and the spectrum of the perceived risks regarding COVID 19 across the six geopolitical zones of the country, with good knowledge translating to a higher perception of risks associated with COVID 19 and poor knowledge otherwise. This finding is similar to what was reported in a global study done on global risk perceptions of COVID 19 in 10 countries across the continents of Europe, Asia and America.^38^ Although in Nigeria about a third of the population are of the opinion that government is doing enough to protect them and their family, about the same fraction feel otherwise with the remaining 33% of the respondents being indifferent to government efforts at ameliorating the effects of COVID 19. This may explain why a significant percentage of the respondents may have adopted for themselves the responsibility of ensuring their safety rather than wait on the government to provide protection for the general populace. These measures being used by individuals may not provide adequate protection against transmission of the disease but rather promote its spread. These are issues which the government agencies task with managing COVID 19 and its complications must investigate. The citizens’ response to safety protocols must be consider critically if any major progress is to be seen in curbing the spread of COVID 19, particularly with the paucity of vaccines in Africa and the possibility of the 3rd wave of the disease already being given serious consideration in other parts of the world.^41,42^

This study has identified good knowledge of the disease and immunization against the virus as the two most important predictors of positive risk perception on COVID 19. This may imply that advocacy on widespread education about the virus and the need for immunization should be embarked upon by the government should any meaningful progress be expected in the fight against the deadly virus.

### Limitations

Findings may be influenced by selection bias because respondents needed access to a smartphone or computer. This may have excluded the poor, elderly who are most vulnerable to COVID-19 this may limit external validity and may have distorted estimation of those willing to take the vaccine.

## Data Availability

Data privacy was ensured

## RECOMMENDATION

It will be imperative for government and non-governmental organizations to strengthen the capacities of health workers to educate and supply information on COVID 19 to the populace because in this research respondents who are health workers tend to have better knowledge of COVID 19 than respondent who are not.

## DECLARATIONS

### Ethics approval

Ethical approval was obtained from the Health Research Ethics Committee of Federal Medical Center, Gusau, Zamfara State, Nigeria

### Consent for publication

Not applicable

### Availability of data and materials

The datasets used and/or analyzed during the current study are available from the corresponding author on reasonable request

### Authors’ Contributions

Conception/design of the study-COO, VKS, OBF, AOO; data collection-UOA, CMI, JCO, OFA, KWO; data analysis and interpretation-EEA, GOP, AOA, OEA; article drafting-BFU, YBA, OFA, AOO; Critical revision of the article-COO, BFU, VKS; final approval of the version to be published-all authors

## Acknowledgment

Oluwafunmike Ruth Olomofe is appreciated for proof-reading the final manuscript.

